# Deep-learning-derived neuroimaging biomarkers of sarcopenia as predictors of outcome in endovascular thrombectomy in large vessel occlusion acute ischemic stroke

**DOI:** 10.1101/2024.11.19.24317593

**Authors:** Kevin Soon Hwee Teo, Benjamin YQ Tan, Yao Neng Teo, Yichi Zhang, Yilei Wu, Yao Hao Teo, Xi Zhen Low, Peng Wu, Joshua YP Yeo, James T P D Hallinan, Li Feng Tan, Christopher D Anderson, Leonard LL Yeo, Andrew Makmur, Juan Helen Zhou

## Abstract

**Introduction:** Sarcopenia is an emerging marker of biological health and is associated with poor outcomes in many disease states. In this study, we aimed to evaluate the utility of muscle biomarkers in predicting clinical outcomes for patients with large vessel occlusion (LVO) acute ischemic stroke (AIS).

**Methods:** This was a single-center observational cohort study of consecutive patients that underwent endovascular thrombectomy (EVT) for LVO AIS. A deep-learning model was employed to segment and measure the volume, surface area, and maximum thickness of temporalis and sternocleidomastoid (SCM) muscles. The primary outcome was poor functional outcome, defined by an mRS of 3-6 at 3 months post-stroke. Univariable and multivariable logistic regression models were performed to evaluate associations between the muscle biomarkers and outcome measures after adjusting for clinical variables of age, sex, and NIHSS.

**Results:** A total of 297 patients were included. 175 (58.9%) had poor functional outcomes at 3 months post-stroke. For each 10cm³ decrease in SCM volume (SV) and temporalis volume (TV), the odds of poor functional outcome at 3 months post-stroke increased by 34% (OR 0.66, 95% CI 0.52–0.84, p < 0.001) and 18% (OR 0.82, 95% CI 0.73–0.91, p < 0.001), respectively. After adjusting for age, sex and NIHSS, our baseline outcome model yielded an AUC of 0.716. Including sarcopenia biomarkers in the model improved discrimination: SV dichotomized (adjusted OR (aOR) 0.39, 95% CI 0.20-0.74, p-value <0.01, AUC: 0.731), TV dichotomized (aOR 0.51, 95% CI 0.30-0.86, p-value 0.012, AUC: 0.724).

**Conclusion:** Our study identified that temporalis and SCM muscle volumes were independently associated with functional outcomes after EVT for LVO AIS.

## INTRODUCTION

Endovascular thrombectomy (EVT), with or without adjunctive intravenous thrombolysis (IVT), has emerged as an effective modality to restore blood flow to ischemic brain tissue in patients with large vessel occlusion (LVO) acute ischemic stroke (AIS) (1,2). However, only half of the patients undergoing EVT achieve functional independence (FI) at 3 months after stroke (3).

Sarcopenia is defined as age-related loss of skeletal muscle and function (4). Sarcopenia has been found to be a strong, independent prognostic indicator of function, nutritional status, morbidity, and mortality in many disease states, including cardiovascular diseases (5). Pre-stroke sarcopenia has been associated with unfavourable outcomes at 3 months post-stroke, higher National Institute of Health Stroke Scale (NIHSS) scores, and lower rates of discharge home post-stroke (6). The measurement of skeletal muscle mass is required for a confirmatory diagnosis of sarcopenia, with the gold standard being computed tomography (CT) or magnetic resonance imaging (MRI) assessment of the hip, thigh, or truncal muscles (7). However, conventional methods to evaluate sarcopenia prove challenging to pursue in the hyperacute stroke situation due to time constraints and limited patient cooperation. The temporalis and sternocleidomastoid (SCM) muscles have emerged as surrogates for skeletal muscle mass measurements for the diagnosis of sarcopenia (8,9), and indices from these muscles have been identified as predictive biomarkers for clinically relevant outcomes in patients with cancer and both hemorrhagic and ischemic stroke (10–12). Previously, studies that segmented head and neck muscles on CT scans primarily adopted atlas-based auto-segmentation methods which can be time consuming and may require substantial manual editing (13, 14). In contrast, employing a deep-learning model may expedite this process through automation, allowing atlas-free segmentation of muscles of interest rapidly and with high fidelity (15).

In this study, we used a deep-learning segmentation model trained on data from a comprehensive stroke center to assess measurements from the SCM and temporalis muscles, readily available for segmentation on an AIS patient’s index CT scan, and their value in predicting clinical outcomes for patients undergoing EVT. We hypothesize that several muscle indices from the temporalis and SCM muscles - muscle thickness, volume, and surface area - may be viable biomarkers in predicting functional independence at 3 months post stroke.

## METHODS

### Study design

This was an observational cohort study of consecutive ischemic stroke patients who underwent EVT for LVO AIS from a single comprehensive stroke center over a four-year period from 2016-2019. LVO was defined as an occlusion of the internal carotid artery, basilar artery, or proximal segments of the middle cerebral artery (M1, M2) and anterior cerebral artery (A1). All patients above the age of 18 who underwent EVT for LVO AIS identified on multidetector CT angiogram (CTA) were included. Patients underwent non-enhanced CT and high-resolution CT angiography as part of their hyperacute stroke assessment. The CT scans were performed on a 64-slice multidetector helical scanner (Philips Inc.) with a 60-70-mL bolus injection of Iohexol contrast. Scan parameters were: source axial thickness of 0.625mm, matrix 512×512, and 120kV. The scan coverage was from the aortic arch to the vertex, and the source images were reformatted into 10×2.5mm axial, coronal, and sagittal maximum intensity projection (MIP) images.

The selection criteria for patients for patients undergoing EVT were in accordance with the American Heart Association / American Stroke Association guidelines for the early management of patients with AIS (16).The exclusion criteria were: patients with hemorrhagic stroke at presentation; altered head and neck muscle anatomy due to previous surgical interventions; neuroimaging with excessive imaging artifact, unusual head posture, and/or insufficient coverage of the SCM or temporalis muscles.

### Data collection

Clinical variables and outcome measures, including age, sex, height, weight, body mass index, comorbidities, NIHSS scores, in-hospital mortality, symptomatic intracranial hemorrhage (sICH) and modified Rankin scale (mRS) scores at 3 months post-stroke were collected. Patients were dichotomized into two categories based on their mRS scores at 3 months post-stroke: those with mRS scores of 0-2 were categorized as attaining functional independence (FI), whereas those with mRS scores of 3-6 were categorized as having poor functional outcomes.

### Manual Annotation

Annotation of patients’ temporalis and sternocleidomastoid muscles was performed using the Darwin Platform software (https://darwin.v7labs.com). All CT annotations were independently performed by two annotators (one consultant neurologist and one consultant radiologist - 4 years’ experience each) and subsequently vetted by board-certified consultant

neuroradiologists (7 to 11 years’ experience) (Supplementary Material). All scan data was anonymized to ensure annotators were blinded to the patient’s comorbidities and outcomes post-EVT.

### Segmentation model training and evaluation

Segmentation model training was carried out using the nnU-Net segmentation framework (15) (Supplementary Material). This enabled networks to be trained to segment three tissue masks - globe, temporalis, and SCM. Three measurements, muscle volume (V), muscle surface area (SA) and muscle maximum thickness (MT) were calculated based on muscle masks with the final measurement an average of the left and right sides (Supplementary Material). For temporalis, these three measurements are abbreviated as temporalis volume (TV), temporalis surface area (TSA) and temporalis muscle maximum thickness (TMT) while for SCM they are abbreviated as SCM volume (SV), SCM surface area (SSA) and SCM maximum muscle thickness (SMT).

To evaluate the performance of the segmentation model, five-fold cross-validation was performed (Supplementary Material). A comparison of similarity between predicted masks and ground-truth labels was assessed using the Dice score, Hausdorff distance (HD) and Absolute Percentage Volume Difference (AVD).

### Outcome measures

The primary outcome measure was poor functional outcome, defined as an mRS score of 3-6 at 3 months post-stroke. Secondary outcome measures were in-hospital mortality and sICH, as defined by ECASS2 consensus criteria (17).

### Statistical analyses

Frequencies and percentages were used to summarize categorical variables and the Pearson’s χ^2^ test for independence was used to compare variables between patients with the primary outcome and those without. Continuous variables following normal distributions were presented as means with standard deviations and compared using the two sample T test; variables that did not follow normal distributions were summarized as medians with inter-quartile ranges (IQR) and compared using the non-parametric Wilcoxon Sign-ranked test.

Univariable regression was performed to identify significant covariates with the primary and secondary outcomes. Multivariable models were built using baseline covariates with statistically significant (p<0.05) associations with the primary and secondary outcomes, including and excluding biomarkers. Odds ratios (OR) were calculated for both univariable and multivariable models.

Muscle parameters were dichotomized for sarcopenia assessment in our cohort of stroke patients. Optimal cutoffs, used to dichotomize sarcopenic biomarkers, were obtained via the Youden’s Index, and area under the receiver operating characteristic curve (AUC) was obtained for the multivariate models. The summary receiver operating characteristic (ROC) curves were subsequently computed and presented in graphs for comparison between the models with and without the included biomarkers. The decision to dichotomize was based on several methodological and clinical considerations. Firstly, the Youden’s Index is a widely recognized method for determining optimal cutoff points in diagnostic tests, as it maximizes the sum of sensitivity and specificity. This allows for a balance between true positive and true negative rates, which is crucial in clinical decision-making. Secondly, dichotomization simplifies the interpretation of results, allowing for a clear distinction between patients with high and low levels of the muscle biomarker, which is particularly valuable in clinical settings where direct categorizations can inform treatment decisions. In the context of stroke patients, where functional outcomes are of paramount importance, a binary classification of muscle biomarkers may provide a more direct link to functional prognosis.

Descriptive statistics and regression models were performed using the R software (version 3.5.2, 2018-12-20; The R Project for Statistical Computing, Vienna, Austria). A statistically significant finding was indicated by a two-sided p value of <0.05.

The study protocol was reviewed and approved by our institutional Domain Specific Review Board. The need for informed consent was waived by the institutional ethics committee and research board as the study design was retrospective and used de-identified patient data that was anonymized prior to extraction by the study teams.

## RESULTS

### Baseline characteristics

The demographic and clinical characteristics of the overall cohort are shown in **Table 1**. A total of 297 patients, with a median age of 68.0 years (IQR 58.0-75.0 years) and 44.4% (n=165) being female, who underwent EVT for large vessel occlusion acute ischemic stroke were included in the study. No patients met the exclusion criteria. FI was achieved by 122 patients (41.1%), while 22 patients (7.4%) developed sICH as a complication. In-hospital mortality occurred in 39 patients (13.1%).

**Table 1.** Baseline characteristics of the study cohort and patients stratified by post-stroke mRS. Abbreviations: ACA - anterior cerebral artery, BMI - body mass index, FI - functional independence (mRS of 0-2 at 3 months), ICA - internal carotid artery, IQR - interquartile range, MCA - middle cerebral artery, mRS - modified Rankin scale, NIHSS - National Institutes of Health Stroke Scale, rTPA - recombinant tissue plasminogen activator, SBP - systolic blood pressure, SCM - sternocleidomastoid, SMT - sternocleidomastoid muscle thickness, SSA - sternocleidomastoid surface area, SV - sternocleidomastoid volume, TMT - temporalis muscle thickness, TSA - temporalis surface area, TV -temporalis volume. Categorical variables presented as n (%) unless otherwise stated. # The TOAST classification denotes five subtypes of ischemic stroke: 1) large-artery atherosclerosis, 2) cardioembolic, 3) small-vessel occlusion, 4) stroke of other determined etiology, and 5) stroke of undetermined etiology.

|  | Overall | FI<br>(mRS 0-2) | Poor functional<br>outcome<br>(mRS 3-6) | p-value |
| --- | --- | --- | --- | --- |
| **n** | 297 | 122 | 175 |  |
| **Age, median [IQR]** | 68.0 [58.0, 75.0] | 63.0 [54.0, 71.0] | 71.0 [62.0, 78.5] | <0.001 |
| **Race** |  |  |  |  |
| **Chinese** | 190 (64.0) | 82 (67.2) | 108 (61.7) |  |
| **Indian** | 25 (8.4) | 9 (7.4) | 16 (9.1) |  |
| **Malay** | 69 (23.2) | 25 (20.5) | 44 (25.1) |  |
| **Others** | 13 (4.3) | 6 (4.9) | 7 (4.0) |  |
| **Sex (% female)** | 165 (44.4) | 73 (40.2) | 92 (47.4) | 0.262 |
| **BMI, median [IQR]** | 24.90 [22.24,<br>27.70] | 25.30 [22.27,<br>27.85] | 24.90 [22.23,<br>27.64] | 0.592 |
| **Hypertension** | 219 (75.0) | 81 (67.5) | 138 (80.2) | 0.02 |
| **Diabetes mellitus** | 92 (31.6) | 30 (25.0) | 62 (36.3) | 0.057 |
| **Hyperlipidemia** | 174 (59.6) | 67 (55.8) | 107 (62.2) | 0.331 |
| **Ischemic heart disease** | 67 (22.9) | 24 (20.0) | 43 (25.0) | 0.391 |
| **Smoking** | 41 (14.2) | 19 (16.0) | 22 (12.9) | 0.579 |
| **Atrial fibrillation** | 131 (44.9) | 50 (41.7) | 81 (47.1) | 0.425 |
| **Previous stroke** | 47 (16.2) | 12 (10.0) | 35 (20.5) | 0.026 |
| **SBP on arrival (mmHg),**<br>**median [IQR]** | 151 [135, 173] | 151 [134, 168] | 152 [136, 176] | 0.371 |
| **NIHSS, median [IQR]** | 19 [13, 23] | 15 [11, 20] | 20 [16, 24] | <0.001 |
| **Thrombolysis with rTPA** | 115 (38.7) | 48 (39.3) | 67 (38.3) | 0.950 |
| **TOAST Classification<sup>#</sup>** |  |  |  | 0.659 |
| **Large artery atherosclerosis** | 66 (22.2) | 25 (20.5) | 41 (23.4) |  |
| **Cardioembolic** | 163 (54.8) | 67 (54.9) | 96 (54.9) |  |
| **Other determined cause** | 9 (3.0) | 5 (4.1) | 4 (2.3) |  |
| **Cryptogenic** | 59 (19.9) | 25 (20.5) | 34 (19.4) |  |
| **Occlusion site** |  |  |  |  |
| **ICA and terminal ICA** | 17 (5.7) | 5 (4.1) | 12 (6.8) | 0.884 |
| **MCA (M1, M2)** | 210 (70.7) | 97 (79.5) | 113 (64.6) | 0.005 |
| **ACA (A1)** | 2 (0.7) | 1 (0.8) | 1 (0.6) | 1 |
| **Basilar artery** | 49 (16.4) | 14 (11.5) | 35(20.0) | 0.088 |
| **Tandem occlusion** | 19 (6.4) | 5 (4.1) | 14 (8.0) | 0.856 |
| **Biomarkers, median [IQR]** |  |  |  |  |
| **SV (cm<sup>3</sup>)** | 26.86 [20.97,<br>35.57] | 30.69 [23.53,<br>38.76] | 25.46 [19.83,<br>34.03] | <0.001 |
| **SSA (cm<sup>2</sup>)** | 79.54 [63.98,<br>92.25] | 83.03 [66.95,<br>94.05] | 74.82 [62.26,<br>90.55] | 0.005 |
| **SMT (mm), mean (SD)** | 13.60 (2.38) | 13.95 (2.28) | 13.35 (2.42) | 0.034 |
| **TV (cm<sup>3</sup>)** | 7.26 [6.08, 8.79] | 8.05 [6.34, 9.18] | 6.94 [5.93, 8.27] | <0.001 |
| **TSA (cm<sup>2</sup>)** | 22.87 [19.35, 26.84] | 24.00 [19.53, 28.09] | 22.19 [19.26, 26.30] | 0.063 |
| **TMT 4mm above globe (mm)** | 7.68 [6.47, 9.43] | 8.39 [6.84, 9.97] | 7.44 [6.33, 8.86] | 0.002 |
| **TMT 6mm above globe (mm)** | 7.27 [6.12, 8.80] | 8.09 [6.57, 9.41] | 6.95 [5.94, 8.37] | 0.001 |

Patients who had poor functional outcomes were older (median 71.0 years [IQR 62.0-78.5] vs. 63.0 years [IQR 54.0-71.0], p < 0.001), had higher NIHSS (median 20 [IQR 16-24] vs. 15 [IQR 11-20], p < 0.001), and had smaller median temporalis volumes (6.94 cm³ [IQR 5.93-8.27] vs. 8.05 cm³ [IQR 6.34-9.18], p < 0.001) as well as SCM volumes (25.46 cm³ [IQR 19.83-34.03] vs. 30.69 cm³ [IQR 23.53-38.76], p < 0.001) compared to those who attained FI.

### Deep-learning-based segmentation performance

The Dice score, which evaluates the spatial overlap between predicted muscle tissue maps and ground-truth manual annotations, were 0.913, 0.950, and 0.880 for the SCM, globe, and temporalis respectively. HD and AVD indices indicated small boundary and volume gaps between the automated and manual segmentation masks (Supplementary Table 2). In Figure 1, the comparison of the predicted muscle segmentation masks and ground-truth manual annotation for two subjects is demonstrated.

**Figure 1.**
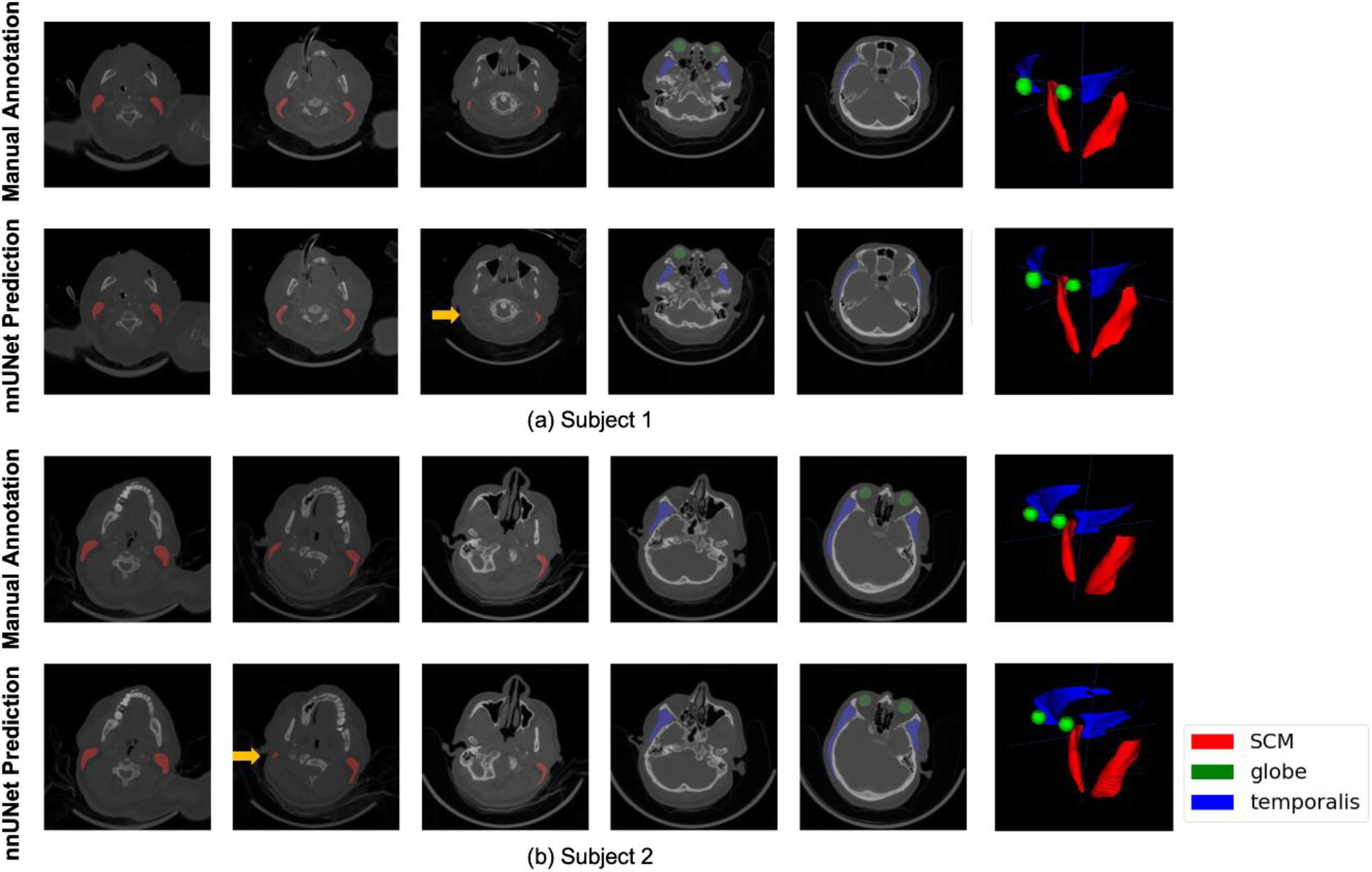
Qualitative visualization of manual and deep-learning model muscle segmentations for two sample subjects. 5 manually annotated and model predicted axial slices are plotted in the first and second row of each panel (green: globe; blue: temporalis; red: SCM), with 3D visualization of the segmented tissues on the rightmost panel. Small tissues on both ends may not be easily detected by the model as indicated by the orange arrows. Abbreviations: SCM - sternocleidomastoid.

### Primary outcome

On univariable analysis, decreased muscle volumes, thickness, and surface areas were associated with higher odds of poor functional outcome at 3 months post-stroke (Table 2). For each 10cm³ decrease in SV and TV, the odds of poor functional outcome at 3 months post-stroke increased by 34% (OR 0.66, 95% CI 0.52–0.84, p < 0.001) and 18% (OR 0.82, 95% CI 0.73–0.91, p < 0.001), respectively. Similarly, for every 1mm decrease in SMT and TMT, the odds of poor functional outcome at 3 months post-stroke increased by 10% (OR 0.90, 95% CI 0.81–0.99, p = 0.035) and 16% (OR 0.84, 95% CI 0.75–0.94, p = 0.003), respectively. Additionally, the odds of poor functional outcome increased by 17% (odds ratio 0.83, 95% CI 0.72–0.94, p < 0.01) for every 1cm² decrease in SSA.

**Table 2.**
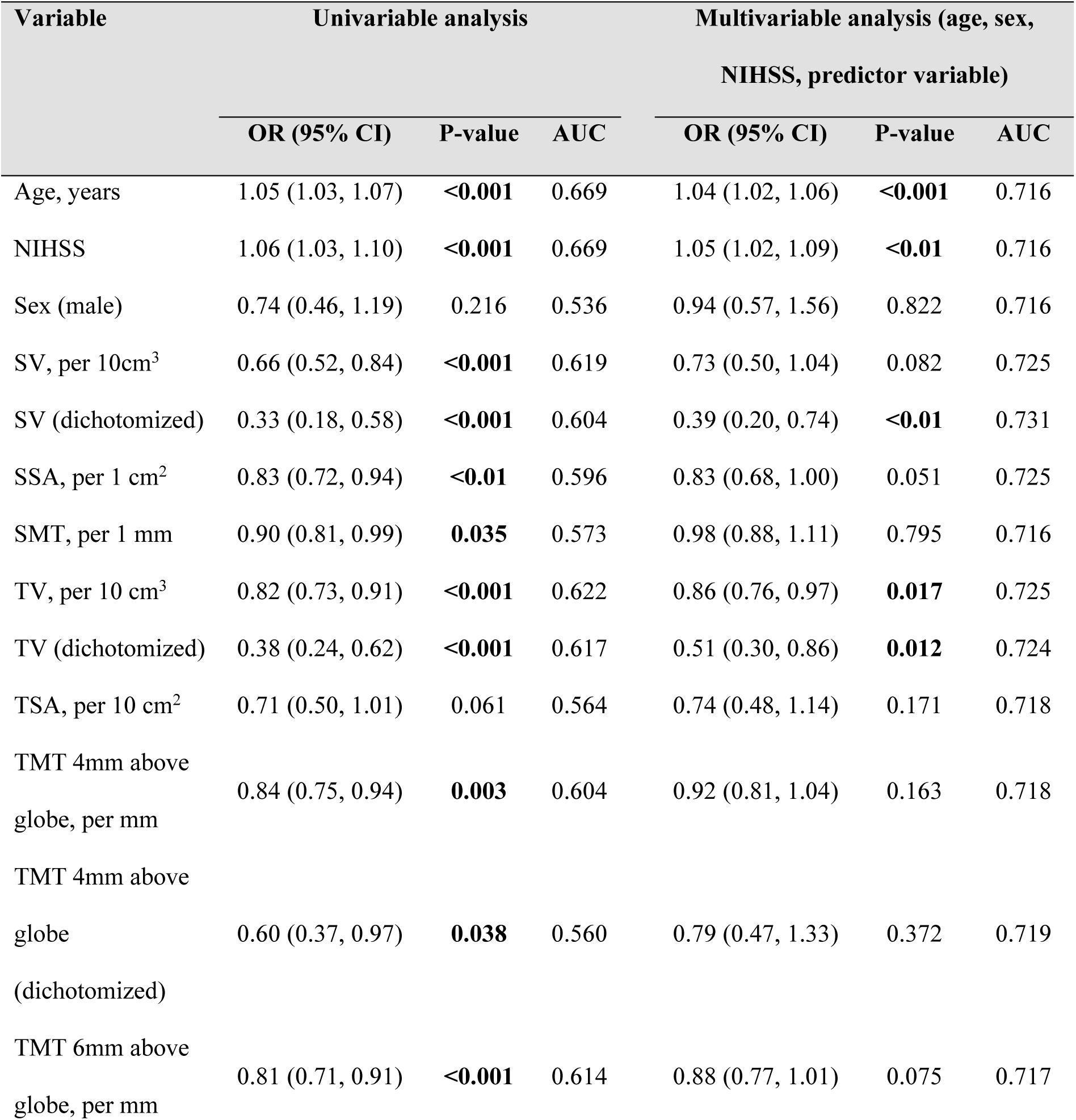
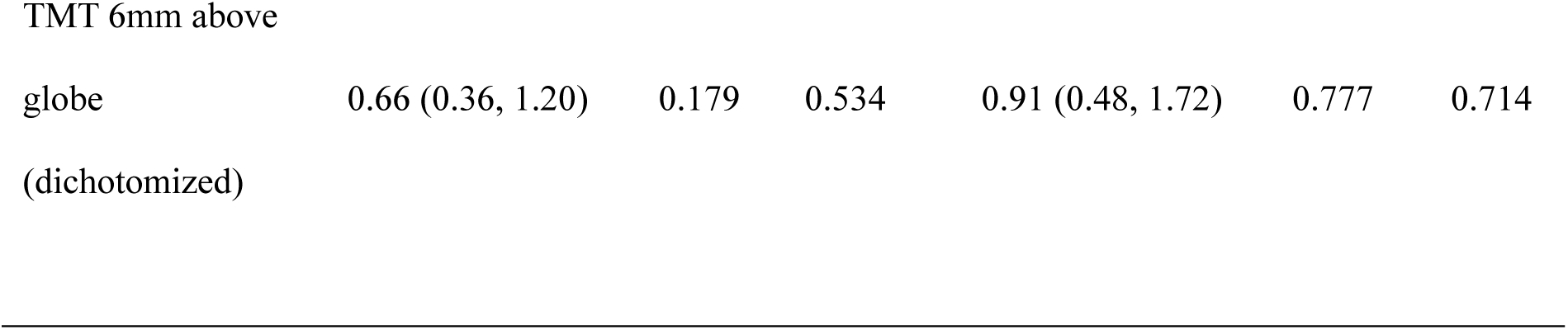
Univariable and multivariable logistic regression models assessing the association between muscle biomarkers and poor functional outcome (mRS 3-6 at 3 months post-stroke) in patients who underwent endovascular thrombectomy for large vessel occlusion acute ischemic stroke. Abbreviations: AUC - area under the receiver operating characteristic curve, BMI - body mass index, CI - confidence interval, NIHSS - National Institutes of Health Stroke Scale, OR - odds ratio, SMT - sternocleidomastoid muscle maximum thickness, SSA -sternocleidomastoid muscle surface area, SV - sternocleidomastoid muscle volume, TMT - temporalis muscle maximum thickness, TSA - temporalis muscle surface area, TV - temporalis muscle volume.

Multivariable models were constructed based on clinical relevance and statistical significance (**Table 2**). Optimal cutoffs for sarcopenia biomarkers, along with Youden’s index and the area under the receiver operating characteristic curve (AUC) at these cutoffs, are presented in **Supplementary Table 4**. Adjusting for age, sex, and NIHSS, the model yielded an AUC of 0.716. Including sarcopenia biomarkers in the model improved the discriminatory ability: SV dichotomized (adjusted OR (aOR) 0.39, 95% CI 0.20-0.74, p-value <0.01, AUC: 0.731, **Supplementary Figure 3**), TV (aOR 0.86, 95% CI 0.76-0.97, p-value 0.017, AUC: 0.725, **Supplementary Figure 4**), TV dichotomized (aOR 0.51, 95% CI 0.30-0.86, p-value 0.012, AUC: 0.724).

### Secondary outcomes

For every 10cm^3^ decrease in SV, the odds of in-hospital mortality increase by 31% (OR 0.69, 95% CI 0.47-0.98, p-value = 0.045). For every 1mm decrease in SMT, the odds of in-hospital mortality increased by 15% (OR 0.85, 95% CI 0.72-0.98, p-value = 0.031) (**Supplementary Table 3**).

In a multivariable model adjusting for age, sex, NIHSS, and sICH, lower SV was associated with an increased odds of in-hospital mortality (aOR 0.47, 95% CI 0.27-0.81, p-value <0.01) post EVT. The model demonstrated fair discriminatory ability (AUC 0.776). No muscle biomarker was significantly associated with sICH on univariable nor multivariable regression.

## DISCUSSION

Our study identified both sternocleidomastoid muscle volume and temporalis muscle volume as significant biomarkers associated with functional independence at 3 months following endovascular therapy for large vessel occlusion acute ischemic stroke. Additionally, sternocleidomastoid muscle volume was associated with in-hospital mortality post-EVT. These muscle biomarkers remained significant predictors of their respective outcomes even after adjusting for age, sex, and NIHSS.

CT assessment is considered the gold standard for non-invasive assessment of muscle quantity and quality and plays a confirmatory role in the diagnostic algorithm for sarcopenia (6). Traditional anatomical locations for skeletal muscle measurements using CT include the thigh, proximal femur, and the L3 lumbar vertebral level (18). There are few studies on sarcopenia as a predictor of outcomes in AIS, which may be due to the difficulty in assessing sarcopenia in AIS patients. For example, the assessment of physical performance and muscle strength may not be feasible in patients who present with weakness. Even in stroke patients who can ambulate, the impact of paresis on these performance measures would limit the validity and reliability of such tests as a tool to assess sarcopenia (6). Nevertheless, opportunistic sampling of temporalis and SCM parameters on routine CT imaging during the hyperacute stroke phase may serve as an invaluable quantifiable imaging biomarker for predicting clinical outcomes in AIS following EVT.

Swartz *et al.* pioneered a method to evaluate skeletal muscle mass using a single CT slice at the level of the third cervical vertebra (19). Their method, which uses defined Hounsfield unit ranges to delineate muscles, has been primarily validated as an alternative method for the diagnosis of sarcopenia in patients with head and neck cancers, whose scans are routinely performed as part of clinical care (11, 20). Since then, TMT, measured using the orbital roof as a common reference point, has also gained popularity as a surrogate marker for skeletal muscle volume, function, and as a prognostic factor in diseases of the head and neck (10–11), with a study by Leitner *et al*. in 2018 demonstrating a high correlation between temporal muscle thickness with standard assessments of lumbar skeletal muscle cross-sectional areas in a cohort of patients with cancer (9). A preceding study by Sakai *et al.* identified TMT as an independent risk factor for post-stroke dysphagia (21). In our study, we utilized the mRS, which is a clinician-reported measure of global disability that has as broad evidence of validity, reliability, and sensitivity as a primary outcome measure in AIS (22). Another study by Nozoe *et al*. found that TMT was not associated with functional independence in a cohort of patients with acute stroke (23). However, this study recruited patients with ischemic and hemorrhagic stroke and comprised a cohort of patients with low NIHSS (mean NIHSS of 3), excluding patients with impaired consciousness, aphasia, or cognitive dysfunction. Consequently, the impact of sarcopenia on functional independence may not have been as significant as it would be in patients with LVO AIS.

More recently, a study by Lin *et al.* identified a significant correlation between TMT and FI at 90 days for patients receiving EVT for LVO AIS, with multivariate analyses identifying TMT as an independent predictor of FI (24). In our study, TMT at 4mm or 6mm above the glove was significantly associated with FI only on univariable analysis but did not achieve statistical significance on multivariable analysis when accounting for age, NIHSS, and sex. Notably, the mean TMT values in our cohort (7.52mm and 7.97mm at 4mm and 6mm above the glove respectively) was comparable to previously published age-group-specific TMT reference values obtained using magnetic resonance imaging in healthy Caucasian volunteers (25), with Lin *et al.* reporting lower mean values for their cohort (6.35mm) compared with reference values. A varying degree of sarcopenia between cohorts may thus have underpinned the discrepancy in results.

A strength of this study was the application of an automated deep-learning model to predict tissue masks on raw cross-sectional CT images. This enabled a time-efficient assessment of various established and potential quantitative CT-derived biomarkers for sarcopenia such as the temporalis and SCM muscle thickness, volume, and surface area and their value in predicting clinical outcomes for patients undergoing EVT for LVO AIS. HD and AVD indices indicate small boundary and volume gaps between the automated and manual segmentation masks. The Dice scores, which identified an 88.0 to 95.0% overlap between manually annotated masks and those generated by a deep-learning approach, indicate a high fidelity which may justify use of our segmentation model in clinical settings or prospective studies which utilize CT imaging of the head and neck as part of disease evaluation or follow-up.

## LIMITATIONS

Large multi-center prospective studies would be required to validate our results as this study was conducted at a single comprehensive stroke center, potentially introducing selection biases. Another limitation was the heterogeneity in the study population, which included patients of multiple ethnicities and included patients with anterior and posterior circulation LVO AIS. Nevertheless, this allowed the study to evaluate EVT outcomes in a real-world setting. Finally, a potential limitation of our study is the lack of normative data of the quantitative CT-derived biomarkers. Normative data provides a baseline reference, allowing for the comparison of individual measurements against a standard population. Without such data, it becomes challenging to accurately interpret the significance of our findings, as we cannot definitively establish whether observed biomarker levels are truly abnormal or within the expected range for the general population. This limitation underscores the necessity for further research to establish normative values within different populations, which would enhance the reliability and clinical applicability of our results. Incorporating normative data in future studies would enable more precise stratification of patient risk and improve the robustness of biomarker-based predictions in acute ischemic stroke.

## CONCLUSION

In our cohort of LVO AIS, temporalis and SCM volumes were found to be independently associated with functional independence at 3 months post-EVT. By applying deep-learning models and relevant neuroimaging biomarkers, clinicians may be able rapidly assess and better predict outcomes of interest for hyperacute stroke revascularization therapy and many diseases of the head and neck.

## Data Availability

The data that support the findings of this study are available from the corresponding author, BYT, upon reasonable request.

## Notes

### Competing Interest Statement

The authors have declared no competing interest.

### Clinical Trial

Not applicable

### Funding Statement

Leonard LL Yeo was supported by the Transition Award, National Medical Research Council Singapore. Benjamin YQ Tan was supported by the National Medical Research Council (NMRC/RTF23jul-0014) and the National University Health System, Singapore (NCSP2.0/2023/NUHS/BTYQ). Juan Helen Zhou was supported by National Medical Research Council and Ministry of Education, Singapore (NMRC/MOH-00707-01; NMRC/OFLCG19May-0035; NMRC/CIRG18Nov-0048) and Yong Loo Lin School of Medicine Research Core Funding, National University of Singapore, Singapore.

### Author Declarations

The study protocol was reviewed and approved by National Healthcare Group (NHG) Domain Specific Review Board, Reference Number 2022/00109. The need for informed consent was waived by the institutional ethics committee and research board (NHG Domain Specific Review Board Reference Number 2022/00109) as the study design was retrospective and used de-identified patient data that was anonymized prior to extraction by the study teams.

